# Forecast Intervals for Infectious Disease Models

**DOI:** 10.1101/2022.04.29.22274494

**Authors:** Rick Picard, Dave Osthus

**Affiliations:** Statistics Group, Los Alamos National Laboratory

**Keywords:** Miscalibration, Overconfident forecasts, Predictive p-values, Predictive tail probabilities, Probabilistic forecasting

## Abstract

Forecast intervals for infectious disease transmission and mortality have long been overconfident — i.e., the advertised coverage probabilities of those intervals fell short of their subsequent performances. Further, there was no apparent relation between how good models claimed to be (as measured by their purported forecast uncertainties) and how good the models really were (as measured by their actual forecast errors). The main cause of this problem lies in the misapplication of textbook methods for uncertainty quantification. A solution lies in the creative use of predictive tail probabilities to obtain valid interval coverages. This approach is compatible with all probabilistic predictive models whose forecast error behavior does not change “too quickly” over time.

## 1 Introduction

Upon reviewing historical epidemiological models of swine flu and foot-and-mouth disease, Ioannidis, Cripps, and Tanner (2020) concluded that “failure in epidemic forecasting is an old problem.” Similarly, with respect to SARS, H1N1, and Ebola, “only a few studies attempted to predict future cases and [they] were ultimately unsuccessful” (Nadella, Swaminathan, and Subramanian, 2020). The same overconfident fate, where advertised forecast accuracy consistently overestimated actual accuracy, has plagued forecasting of the COVID-19 pandemic.

Prediction overconfidence affects the biological sciences more broadly for many of the same reasons as it does for infectious disease modeling. Numerous examples exist (Ioannidis, 2005) involving drug efficacy and assessment of therapeutic treatments, as well as statistical analyses of fMRI data (Eklund, Nichols, and Knutson, 2015) and microarray data (Michiels, Koscienly, and Hill, 2005). Overconfidence issues are minimized only when there is a clear distinction between exploratory and confirmative data analyses, such as for Phase II and Phase III clinical trials (Tong, 2019).

Most development of complex predictive models involves learn-as-you-go modeling. Models are derived based on goodness of fit checks to available data and are revised over time in light of updated information in a quest for ever improved forecast performance. This model development process often leads to reasonable point prediction but, as detailed in Section 2, also leads to overconfidence in purported forecast accuracy because textbook methods for uncertainty quantification usually fail for this type of modeling. In Section 3, we discuss different types of infectious disease models, and in Section 4 describe how overconfidence manifests itself in them. We then show how to correct for poor forecast intervals through the use of predictive tail probabilities in Section 5.

## 2 Overconfidence: It’s Real and Pervasive

Critiques of epidemiological forecasting have often been anecdotal. A topical example involves COVID-19. Early one-day-ahead death predictions from a prominent model were such that nominal 95% coverage intervals provided only 30% coverage (Begley, 2020). Similar issues with other models led to serious criticism of COVID-19 forecasting (Holmdahl and Buckee, 2020; Ioannidis, Cripps, and Tanner, 2020; Wynants et al., 2020; Recchia, Freeman, and Spiegelhalter, 2021). A corresponding blizzard of negative media coverage (for a small sampling, see Marfin, 2020; Sullum, 2020; Olson, 2021) unsettled image-conscious decisionmakers from all across the political spectrum, leading one frustrated state governor to remark of COVID-19 forecasts that “I’m out of that business because we all failed at that business. Right? All the early national experts. Here’s my projection model. Here’s my projection model. They were all wrong.” (as quoted in Chin et al., 2020, p. 733)

The recent emphasis of data repositories on probabilistic forecasting provides a more detailed understanding of the problem and also offers a path to its solution. Working with the U.S. Centers for Disease Control and Prevention (CDC), the COVID-19 Forecast Hub (Wattanachit et al., 2020) is the central repository for COVID-19 forecasts. Forecasts of future case counts and deaths for U.S. states are submitted to the Hub. Case numbers and deaths are available daily, and forecasts are assessed weekly.

To quantify overconfidence in predictions, one-week-ahead cumulative deaths forecasts are summarized for the 20 COVID-19 models that provided regular submissions for at least 49 states between June 1st, 2020 and March 31st, 2021. Forecasts were in the form of 23 percentiles (1%, 2.5%, 5%, 10%, …, 95%, 97.5%, 99%), and overconfidence was assessed over this time period.

Figure 1 shows the empirical coverage results directly and as overconfidence factors, quantifying the latter as the ratio of tail probabilities

**Figure 1:**
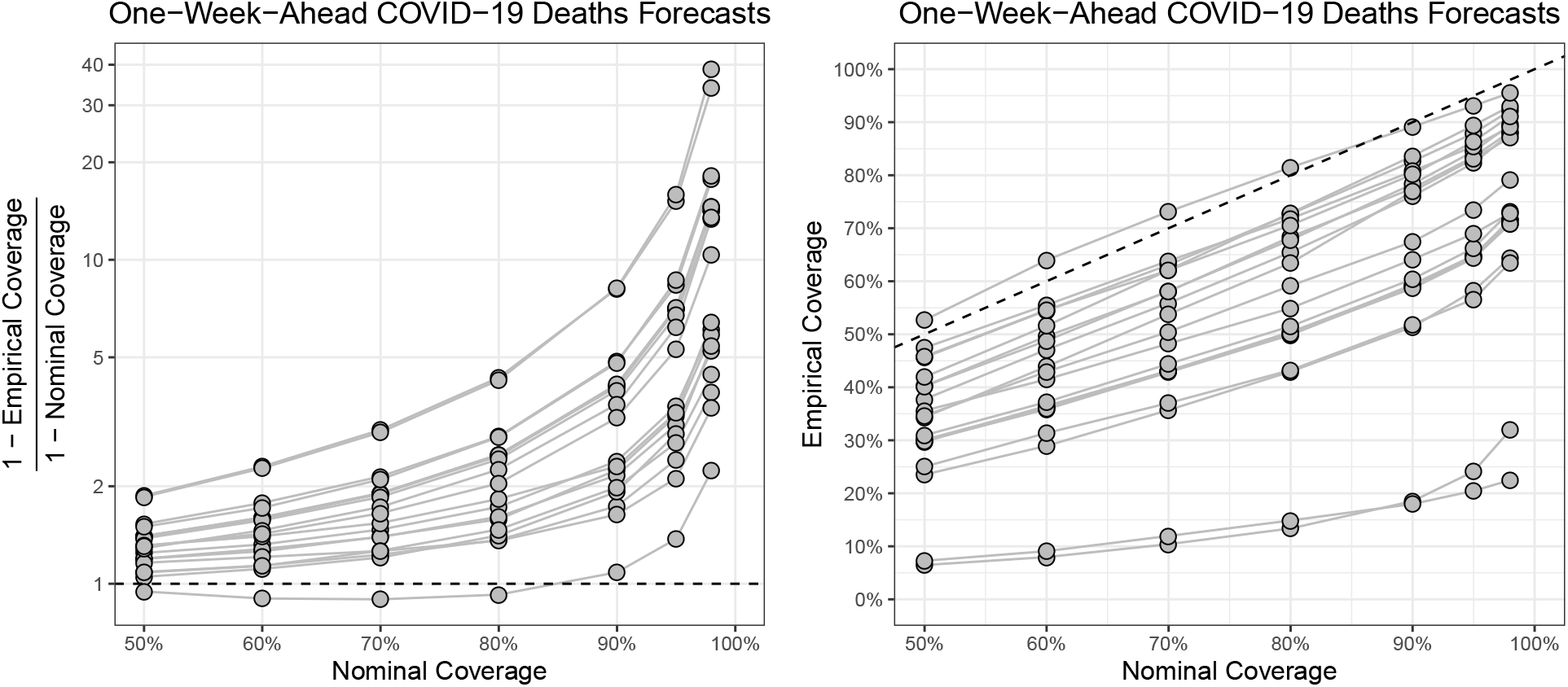
Left: Overconfidence factors for 20 one-week-ahead COVID-19 cumulative death forecast models (in log scale to minimize overplotting). Each segmented line is a forecast model. Right: Empirical versus nominal coverage probabilities.

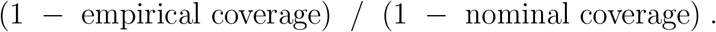

For example, if a nominal 95% forecasting interval provided only 85% coverage, the over-confidence factor 15%*/*5% = 3 indicates that there were three times the expected number of predictions falling outside their nominal prediction intervals. Thus, an overconfidence factor of 1, plus or minus statistical variation, would reflect a well calibrated forecast model.

Figure 1 displays predictive performance across the range of nominal coverage probabilities of greatest interest. It also allows modelers to assess whether the “false positive rate” for a particular coverage-probability-specific interval is of practical concern.

The most striking aspect of Figure 1 is that overconfidence is pervasive. Very few forecast intervals are reasonably well calibrated. Nearly all models generated 95% and 98% forecast intervals for which several times as many observed values fell outside their prediction intervals than would be expected. Overconfidence factors between 3 and 10 were typical, with numerous factors exceeding 10. In that each point in Figure 1 is based on roughly 2000 forecasts (more than 40 weekly forecasts for each of 50 U.S. states), the statistical evidence for overconfidence is overwhelming.

Another aspect of overconfidence addresses the (lack of a) relationship between how well models claim to forecast and how well they actually do, shown in Figure 2. Note that there is no apparent relation between purported and actual COVID-19 forecasting ability, results that run counter to what would ideally be expected.

**Figure 2:**
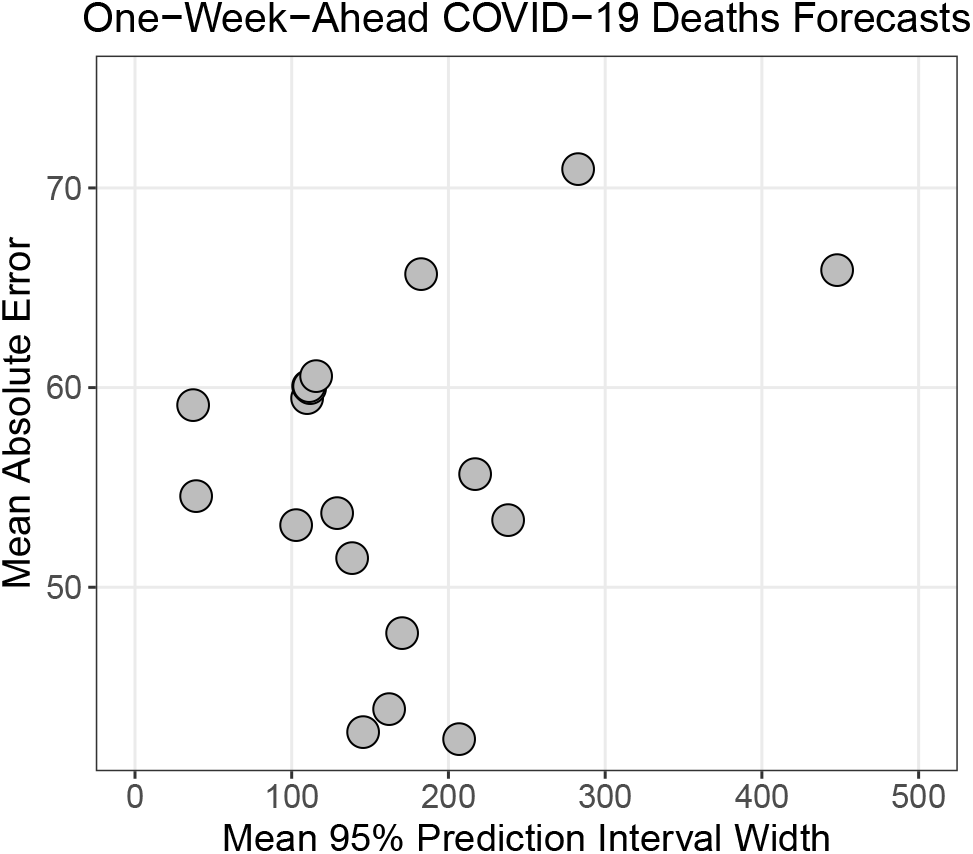
Mean absolute error (MAE) (y-axis) and average 95% prediction interval width for one-week-ahead cumulative COVID-19 deaths forecasts. Each point is a forecast model. One outlier model at MAE = 80 and mean 95% prediction interval width ≈ 2500 (driven by a small number of extremely large 95% prediction interval widths) was removed for improved plotting.

## 3 Predictive Models and Forecast Intervals

Infectious disease models can loosely be categorized as mechanistic, statistical, and ensemble combinations, although the classification between mechanistic and statistical models is sometimes blurred. This distinction among model types is important because different modeling approaches naturally lead to different methods for constructing forecast intervals, which in turn lead to different causes of overconfidence.

Purely mechanistic models attempt to mimic actual disease spread. They do so via various input parameters – such as the basic reproduction number *R*_0_, the mortality rate as dependent on comorbidities, the portion of the population that has either recovered from the disease or has been vaccinated and is believed to be far less susceptible to disease-related death, etc. Depending on the model resolution (agent-based computer models being an extreme example), such input values may be required for several subpopulations of individuals in a delicate balancing act between the potential benefits of increased model resolution and the drawbacks of decreasing input data quality for smaller subpopulations.

The actual values of input parameters for mechanistic models are unknown, difficult to measure directly, dependent on the geographic location(s) involved, and can dynamically respond to changes in societal behavior. Based on the input values, a mechanistic model attempts to quantify the disease progression. Forecast intervals arise upon propagating input uncertainties through a computer code. Upon repeated Monte Carlo sampling of uncertain model inputs and making code runs for each sampled set of inputs, a spectrum of output forecast horizons is obtained and used in the formation of prediction intervals.

Purely statistical models treat observed data streams as nonstationary time series with the goal of future prediction. The number of parameters in such models varies from a few for simple time series and Kalman filter models to many for machine learning approaches. Forecast intervals for statistical models arise from applying standard textbook results.

Lastly, there exist ensemble combinations of individual mechanistic/statistical models which can, in principle, leverage distinct strengths of the respective models to better give results than from any individual model alone. Methods for combination include Bayesian model averaging, frequentist model averaging, and various weighting schemes.

## 4 Overconfidence: The Root Causes

As is apparent from Sections 1 and 2, overconfidence is pervasive. Were a mechanistic model formally correct, or a statistical model valid in terms of its underlying assumptions, overconfidence would not occur. Clearly, the textbook interval determination methods used for infectious disease forecasting are being (mis)applied based on faulty premises.

Model development in learn-as-you-go settings often begins by first considering models that have been used in similar problems, such as considering for COVID-19 models that have been used for influenza-like-illness transmission/mortality. Tentative models are then force-fit to available data. Upon assessing various goodness-of-fit measures, modifications are made as deemed necessary. Importantly, the effects of these modifications — made subjectively by the modeler via so-called “researcher degrees of freedom” (Simmons, Nelson, and Simonsohn, 2011) — are nowhere accounted for in textbook uncertainty analysis.

### 4.1 Purely mechanistic models

The Monte Carlo approach to uncertainty quantification for mechanistic models samples computer model inputs from a perceived joint probability distribution, runs the model with each sampled set of inputs, and then examines the spectrum of output quantities. A shortcoming of this formalized sensitivity analysis, however, is that subject matter experts are notorious (Lin and Bier, 2008) for understating actual uncertainties such as for model parameters. Similarly, any overconfidence in the learn-as-you-go patient-level modeling that helps supply input parameter distributions (such as for subpopulation-specific transmission and mortality rates) naturally leads to overconfident intervals for mechanistic models.

An additional factor is that the full range of human behavior (e.g., for COVID-19, varied societal adherence to ever-evolving government directives, a fatigue that can set in over the long-term) and clinical factors (e.g., continual virus mutation, improved therapeutic treatment over time as more is learned about a disease, effects of widespread vaccination) are difficult to anticipate owing to a lack of historical experience, and are easily underestimated. Consequently, the Monte Carlo approach does not fully sample the input space, and the range of output uncertainty is understated. Further, the use of “projections” (i.e., assuming specific values or ranges for certain input parameters to assess “what if” scenarios for policy planning) only complicates the interpretation of results.

### 4.2 Purely statistical models

Textbook formulas unfortunately do not apply in learn-as-you-go environments. When misused, overconfident prediction intervals are expected (e.g., Picard and Cook, 1984; Chatfield, 1995). Despite such admonitions, the overconfidence phenomenon is not considered in most contemporary epidemiological modeling, as evidenced by pervasively overconfident forecasts.

Classical statistical theory for prediction accuracy assumes that

1. The “known” form of the statistical model is determined independently of the data, and
2. textbook prediction intervals follow from the model being probabilistically correct.

These assumptions do not apply for learn-as-you-do models, especially in regard to the effects of researcher degrees of freedom in model development. Tong (2019) gives an insightful historical review of overconfident statistical inference.

Overconfidence not only affects prediction intervals, but it also affects confidence intervals for model parameters and related significance tests. Indeed, there are strong parallels between the historical misuse of ordinary p-values in data analyses and the current misuse of textbook prediction interval methods for learn-as-you-go epidemiological models. That is, scientists were misled for years to believe apparent data-based p-values provided much greater certainty than was warranted, a problem that had long been recognized by the American Statistical Association (Wasserstein and Lazar, 2016) and statistics community in general. Until the misuse finally spiraled out of control, however, reactions took the form

> “yes, we know about it, yes, we have considered taking action – but no, we are not going to do so because it would cause too much upheaval” (Matthews, 2017, p. 39).

To this point, reactions to learn-as-you-go modeling overconfidence are evolving similarly. Bayesian counterparts of classical statistical models, designed to better leverage available subjective knowledge in addition to the observed data, further presume that

3. The prior distribution on model parameters is divined independently of the data, and
4. the prior distribution fully describes available subjective knowledge.

These assumptions also do not apply for learn-as-you-go modeling. Any dependence of a prior on the data invalidates the usual application of Bayes’ theorem. Moreover, when a range of prior distributions is consistent with available knowledge (as is often the case), restricting attention to a single prior underestimates uncertainty; at a minimum, informal sensitivity analysis for the prior distribution is warranted. Tendencies for Bayesian applications to be “optimistic with their model assumptions” (Efron, 2005, p. 1) and to “discount the possibility that [they] may be miscalibrated” (Dawid, 1982, p. 606) further exacerbate overconfidence. Computationally, incomplete convergence of Markov chain Monte Carlo algorithms can easily occur and contribute to unduly narrow credibility regions, especially when parameter identifiability issues exist (Roda et al., 2020) — e.g., death counts from a highly transmissible but low-lethality mutation can strongly resemble those from a less transmissible but somewhat more lethal mutation.

## 5 Calibrated Forecasts in High-Feedback Environments

Despite the lack of a formal methodology to account for subjective model development, a solution for overconfidence exists when there is a sufficiently high volume of forecasts. Consider, for example, predicting COVID-19 deaths for the 50 U.S. states. As data become available each day, forecast errors are revealed. The one-day-ahead forecast errors for forecasts made yesterday can be computed, as can errors for two-day-ahead forecasts made two days ago, errors for one-week-ahead forecasts made one week ago, etc.

This feedback allows for forecast errors to be assessed relative to their nominal probabilistic forecast distributions. As each such forecast error is observed, compute its predictive tail probability relative to its nominal probabilistic forecast distribution. That is, compute the portion of the probabilistic forecast distribution that exceeds the corresponding observed value that it was predicting. In Bayesian models, this quantity is called a predictive p-value (Rubin, 1984; Gelman, Meng, and Stern, 1996), though we use the catch-all term predictive tail probability to extend this concept to the variety of epidemiological forecasting methods.

Were one-week-ahead death forecasts well calibrated, 95% of the predictive tail probability values would be between 2.5% and 97.5%. From Section 2, forecasts are generally overconfident, and their too-narrow forecast intervals should be widened. Such a widening can be achieved “on the fly” using recently observed predictive tail probabilities, as we soon show.

To be sure, the concept of using observed forecast errors to “calibrate” or “de-bias” poorly calibrated forecast intervals is not new (e.g., Cox, 1958; Graziani et al., 2019; Clemen and Lichtendahl, 2002). The novelty of the approach herein lies in its direct use of predictive tail probabilities, pooling them over non-exchangeable probability distributions to obtain valid intervals in real time across arbitrary forecast error correlation structures. See the next section for theoretical details.

The approach is based on the principle that good interval calibration is produced when:

1. The stochastic forecast error behavior relevant to a “current” day’s predictions is similar to the behavior for recent days’ predictions, and
2. the set of forecasts over which calibration is to be assessed, such as forecasts for the 50 U.S. states, is clearly specified.

Although Assumption 1 cannot be literally true in a forecasting environment as fluid as for infectious diseases, it provides a far better approximation to reality than do standard textbook assumptions, and it leads to greatly improved coverage rates.

In the example to follow, a cursory examination of various window widths deemed as “recent” all forecast errors that became known during the 14 days immediately before a current prediction. The idea is to compile the empirical cumulative distribution function (CDF) of predictive tail probabilities from recent forecasts and use the quantiles of that CDF to define forecast intervals.

Suppose, in the COVID-19 example, that exactly 2.5% of the most recent 700 (14 days and 50 U.S. states per day) predictive tail probability values were less than 0.01 and exactly 97.5% of those values were less than 0.98. Then the 95% forecast intervals for today’s 50 state-specific forecasts would be between the 1% and 98% (upper) quantiles of today’s state-specific probabilistic forecast distributions (in contrast to the too-narrow 95% nominal intervals between the 2.5% and 97.5% quantiles of those same distributions).

Had these empirical values (1% and 98% in this hypothetical example) been omnisciently used for the corresponding 14 days’ forecast intervals, perfect overall calibration of those forecasts would have been achieved. Although it is not possible to omnisciently go back in time, it is pragmatic to use those values for today’s forecasts. So long as the marginal distributions of U.S.-state-specific daily predictive tail probabilities don’t change much from day to day (the formalized version of Assumption 1), valid forecast intervals are obtained.

A potential issue with the approach involves longer-term forecasts. Consider, for example, making two-week-ahead COVID-19 death forecasts for the 50 U.S. states on a daily basis. The 50 two-week-ahead forecast errors observed today correspond to forecasts made 14 days ago; the 700 most recently observed forecast errors correspond to forecasts that were made 14-27 days ago. Assumption 1 above postulates that the two-week-ahead forecast error behavior that is relevant to today’s forecast is similar to that for two-week-ahead forecasts made 14-27 days ago. Were forecast error behavior to change too much, too quickly, calibration would be affected. We revisit this subject in the example of Section 7.

## 6 Theoretical Motivation

The primary challenge to developing theory for prediction intervals is that probability distributions of forecast errors are unknown and unknowable for learn-as-you-go models. Not only are marginal distributions unknown, but so are correlations among distinct forecast errors (e.g., those between one forecast horizon time and another). Thus, valid intervals must be developed without reliance on such knowledge.

To do this, consider a particular forecast, e.g., one-week-ahead cumulative deaths. Probabilistic forecasting is based on a perceived probability distribution for the predicted deaths 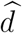. Nominal forecast intervals are defined by quantiles of this distribution.

Once the corresponding death count *d* is observed, its predictive tail probability is the portion of the perceived probability distribution for 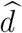 that exceeds *d*. When forecast intervals are well calibrated across all coverage probabilities, predictive tail probabilities are uniformly distributed over (0,1). For overconfident forecasts, their distribution tends to be U-shaped.

Overconfidence is resolvable through calibration. Notationally, let

*P* ^(*i*)^ denote the predictive tail probability for state *i* for the “current” day,

*F* ^(*i*)^(*p*) denote the (unknown) CDF of *P* ^(*i*)^, *p*_0_ denote a constant in (0,1), and

*X*^(*i*)^ denote the indicator function of the event *{P* ^(*i*)^ ≤ *p*_0_*}*.

The portion of states whose predictive tail probabilities for the current day will be less than or equal to *p*_0_ is thus Σ*X*^(*i*)^*/*50, where the sum is over the 50 states. The expected value of the current day’s portion is

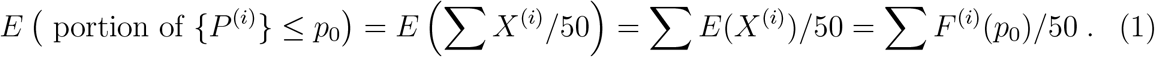

Note that (1) applies for all state-to-state correlation structures among the *{P* ^(*i*)^*}*.

Suppose (just for the moment) that day-to-day independence holds, so that the current day’s 50 U.S.-state-specific predictive tail probabilities together with those from the 14 most recent days’ forecasts, form an independent sample of size 15 from a 50-dimensional distribution. In this case, predictive tail probabilities for the *i*-th state for the current and all 14 recent days have common marginal distribution function *F* ^(*i*)^. For *p*_0_ ∈ (0, 1) as above, and 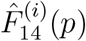 denoting the empirical CDF of predictive tail probabilities for the *i*-th state observed over the 14 most recent days, the collective portion of all 50-state recent predictive tail probabilities less than or equal to *p*_0_ can be written

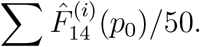

The expected value of this quantity is

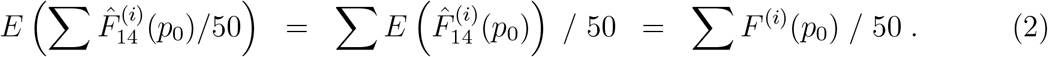

[Aside. The above assumption of day-to-day independence of predictive tail probabilities is not necessary for (2). For the non-independence case, view 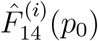 for each *p*_0_ as the average of 14 correlated-but-identically-distributed day-specific indicator functions similar to *X*^(*i*)^ above. Linearly accumulating the expectations through the average as in Equation (1) above yields 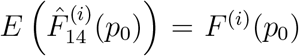. Consequently, the equality of (1) and (2) is not affected by either the day-to-day correlations across recent forecasts or by the state-to-state correlations in the 50-dimensional multivariate distribution for the *{P* ^(*i*)^*}*.]

Returning to the hypothetical example in Section 5, suppose that exactly 2.5% of recent predictive tail probabilities were less than or equal to 0.01 and 97.5% of recent predictive tail probabilities were less than or equal to 0.98, i.e.,

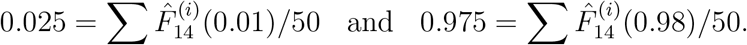

Using these quantiles of the empirical CDF gives the desired result for the current day’s predictive tail probabilities *{P* ^(*i*)^*}*:

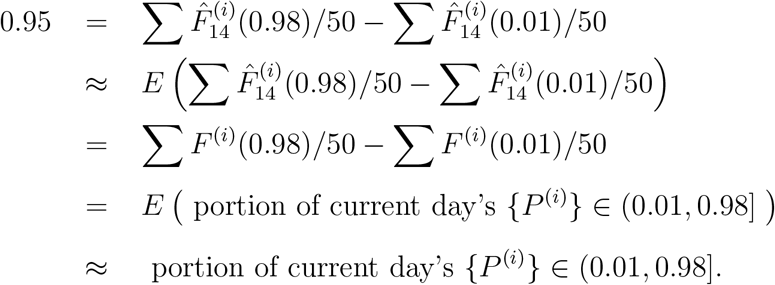

The two approximations in the above should be noted. The first reflects the variation in the recent days’ predictive tail probability quantiles, and the second reflects the variation in the current day’s predictive tail probability quantiles. For the high-feedback COVID-19 data, recent days’ quantiles are based on a (non-independent) collective sample of size 700, which is more than adequate unless extreme tail probabilities are required. The second approximation in (2) reflects that the current day’s quantiles are based on a (similarly non-independent) collective sample of size 50, adequate when assessed over a reasonable time frame to give good overall results, as illustrated in the example to follow.

Finally, the assumption that the marginal distributions *{F* ^(*i*)^*}* of state-specific predictive tail probabilities are exactly the same for the current and all “recent” days is unrealistically constraining. Pragmatically, when the marginal distributions don’t change too much, too quickly over time, recent forecast errors are representative of current ones and good collective calibration of prediction intervals occurs.

## 7 An Example

We illustrate using the forecast model COFFEE: COVID-19 Forecasts using Fast Evaluations and Estimation (Castro et al., 2020). This model is chosen because its complexity is typical of infectious disease models, but it is not overly demanding computationally (so that 10 months of daily 50-U.S.-state-specific forecast updates can be produced in a tolerable CPU time).

COFFEE produces daily forecasts upon combining short-term modeling (an instantaneous daily change rate coupled with a day-of-week effect) with longer-term modeling (anticipating future societal behavior by postulating that if daily deaths have increased for too long, or decreased for too long, the curve will tend to bend in the opposite direction). Researcher degrees of freedom for the COFFEE algorithm are considerable, including: automated outlier identification/adjustment to account for data eccentricities, dynamic variable selection for day-of-week effects, and weights in regressions that are based on inverted influence metrics. These researcher degrees of freedom do not allow for tractable analytical theory. Instead, data splitting (Picard and Berk, 1990) is used in a parameter estimation context.

Based on the data splitting, a joint probability distribution for certain model input parameters, scaled by agreement with test-set data, is derived. Then, similar to the Monte Carlo approach to uncertainty quantification used for mechanistic models, samples from this distribution are obtained, input to the COFFEE model, and samples of future COVID-19 forecasts are simulated. These output samples define the probabilistic forecast distributions used to construct nominal forecast intervals.

Owing to the optimism principle (Picard and Cook, 1984), parameter estimates optimized to current data will not give the same level of performance when used for prediction, and the nominal COFFEE prediction intervals are expected to be overconfident. Results in Figure 3 quantify this effect for one-week-ahead forecasted cumulative deaths.

**Figure 3:**
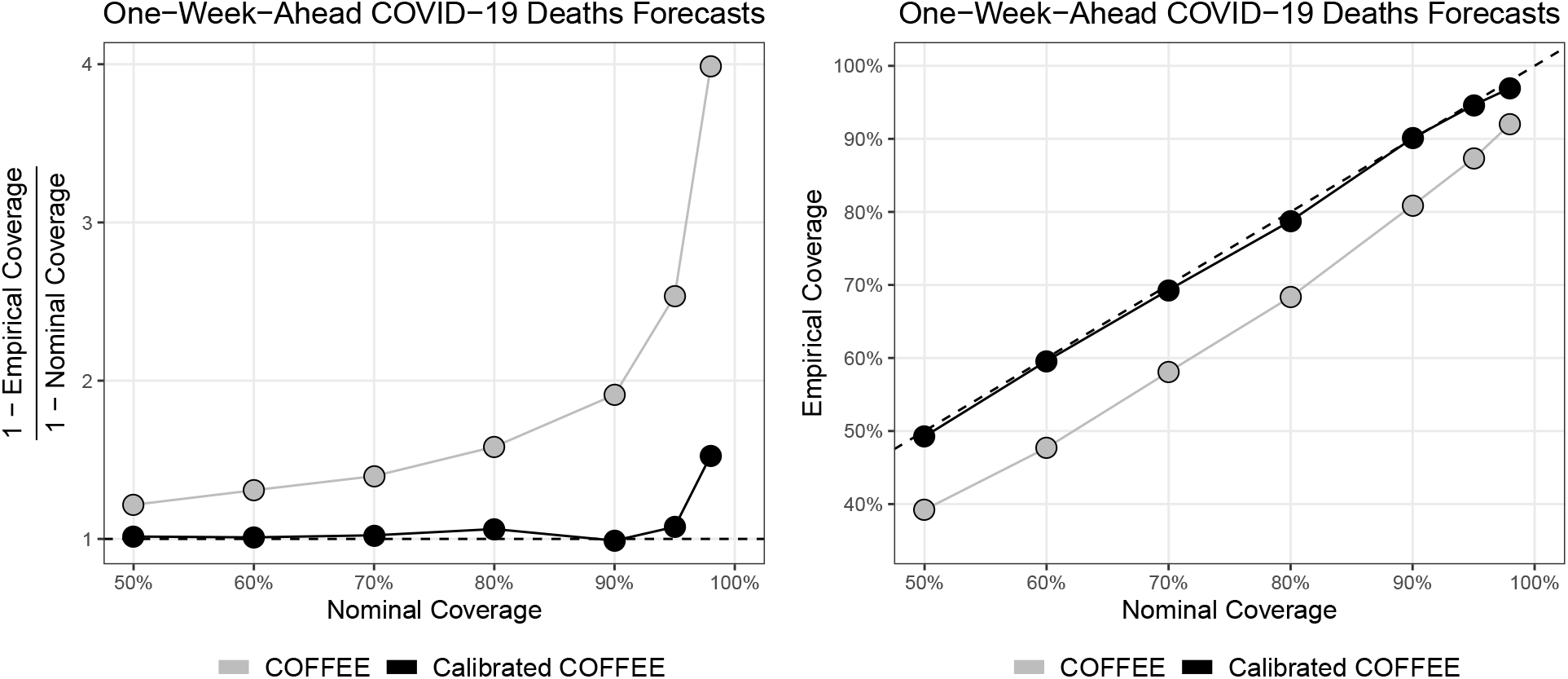
Left: One-week-ahead cumulative death overconfidence factors for original COFFEE forecasts (grey) made between June 1st, 2020 and March 31st, 2021, and calibrated COFFEE forecasts (black). Right: Empirical versus nominal coverage probabilities.

Now replay the data accumulation, calibrating nominal COFFEE forecast intervals on the fly based on predictive tail probabilities from the 14 days immediately preceding each forecasting day. Upon calibration, overconfidence is greatly diminished. See Figure 3.

For longer-term forecasts, the calibration approach does not ensure perfect interval coverages, as shown for 2- and 3-week-ahead forecasts in Figure 4. Overconfidence factors are greatly reduced at the 90%, 95%, and 98% nominal coverage probabilities, but there exists residual overconfidence. This is largely due to recently observed forecast errors at longer time horizons corresponding to forecasts that are not themselves so recent, as noted at the end of Section 5, which undercuts Assumption 1 and limits effectiveness somewhat.

**Figure 4:**
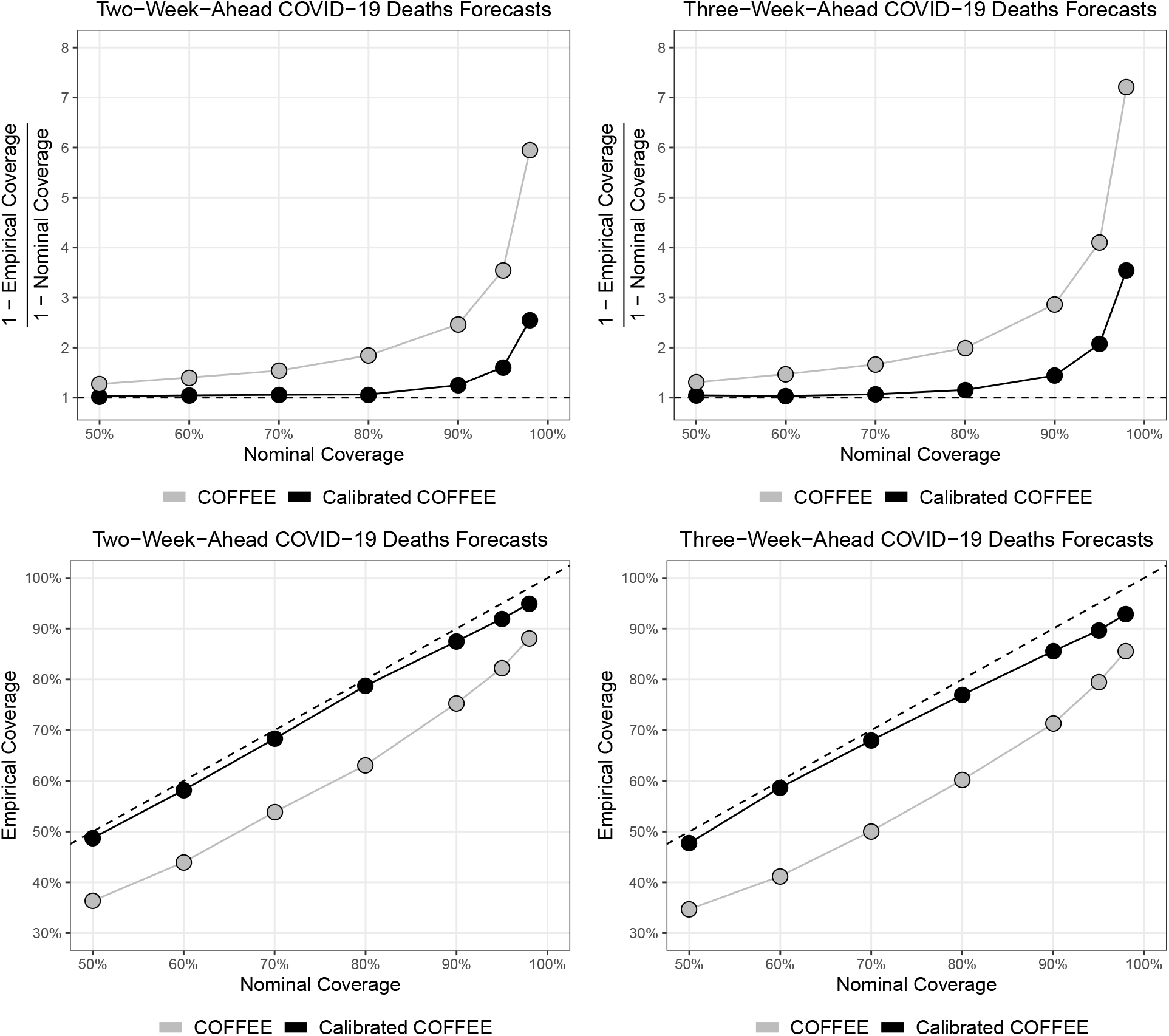
Top: Two- (left) and three-week-ahead (right) cumulative death overconfidence factors for original COFFEE forecasts (grey) and calibrated COFFEE forecasts (black). Bottom: Empirical versus nominal coverage probabilities.

Perfect calibration occurs only when there exists pristine textbook theory underlying a prediction interval method. Force-fitting such pristine theory to learn-as-you-go models where it does not apply leads to severe overconfidence as per Figure 1. Calibration corrects for overconfidence and vastly improves overall coverage probabilities.

## 8 Conclusion

At its best, forecasting provides real-time, reliable, and actionable insights to decision-makers. At its worst, forecasting provides overly-confident and misleading information to decisionmakers. The latter has occurred all too often for infectious disease predictions as discussed in Sections 1 and 2. The calibration approach corrects for overconfidence when high-feedback data exists; even in lower-feedback environments, the lessons described herein regarding the root causes of overconfident prediction are important to modelers.

Specifically, COVID-19 forecast data, accumulated over tens of forecast models, the 50 U.S. states, multiple forecast lead times, and thousands of total forecasts, conclusively showed:

1. Standard textbook methods, when misused in dynamic learn-as-you-go prediction environments, produced pervasively overconfident forecast intervals.
2. Overconfidence factors for COVID-19 worsened as nominal coverage probabilities increased (e.g., from 90% to 95% to 98%); predicted death counts fell outside their nominal prediction intervals by factors of 3-10 times more often than naively expected.
3. Forecast models having small advertised forecast interval widths did not produce any better actual forecasts than did models having larger advertised forecast interval widths.

For high-feedback environments, the problem of poorly calibrated forecast intervals is overcome by using predictive tail probabilities. This approach is compatible with any model, can be implemented in real time with minimal programming effort beyond that already required for probabilistic forecasting, and naturally makes the right adjustments whether the model is overconfident, well calibrated, or ultra conservative. The approach works whenever forecast error behavior does not change too much, too quickly.

Last but by no means least, although we have focused on obtaining valid forecast intervals, the bigger picture warrants attention. Not only should a model’s advertised interval coverages be well calibrated (or “honest”), but the intervals should also be sufficiently narrow (or “sharp”) as to be of practical value to decisionmakers. The ultimate goal for probabilistic forecasting is “maximizing the sharpness of predictive distributions *subject to calibration*” (Gneiting, Balabdaoui, and Raftery, 2007, p. 243, emphasis added). Pursuit of this goal has been hampered because nearly all epidemiological probabilistic forecasting has failed the calibration requirement. So long as only poorly calibrated intervals are available, the true forecasting ability of a model is difficult to determine, leaving decisionmakers in the unenviable position of taking real-time actions without good knowledge of the actual uncertainties involved. As such, formal methodology for producing valid forecast intervals is a significant step towards better decisionmaking.

## Data Availability

Forecast data are accessible from https://covid19forecasthub.org/

## Acknowledgements

We thank Nishant Panda, Isaac Michaud, and Sara Del Valle for graphical, computational, and funding assistance. This work was developed with the support of LANL’s LDRD research Projects 20200698ER and 20190546ECR.

## Notes

### Competing Interest Statement

The authors have declared no competing interest.

### Funding Statement

This work was developed with the support of LANL's Laboratory Directed Research and Development projects 20200698ER and 20190546ECR.

